# Protocol for a bandit-based response adaptive trial to evaluate the effectiveness of brief self-guided digital interventions for reducing psychological distress in university students: The Vibe Up Study

**DOI:** 10.1101/2022.12.05.22283129

**Authors:** Kit Huckvale, Leonard Hoon, Eileen Stech, Jill Newby, Wu-Yi Zheng, Jin Han, Rajesh Vasa, Sunil Gupta, Scott Barnett, Manisha Senadeera, Stuart Cameron, Stefanus Kurniawan, Akash Agarwal, Joost Funke Kupper, Joshua Asbury, David Willie, Alasdair Grant, Henry Cutler, Bonny Parkinson, Antonio Ahumada-Canale, Joanne R Beames, Rena Logothetis, Marya Bautista, Jodie Rosenberg, Artur Shvetcov, Thomas Quinn, Andrew Mackinnon, Santu Rana, Truyen Tran, Simon Rosenbaum, Kon Mouzakis, Aliza Werner-Seidler, Alexis Whitton, Svetha Venkatesh, Helen Christensen

## Abstract

**Introduction:** Meta-analytic evidence confirms a range of interventions, including mindfulness, physical activity and sleep hygiene, can reduce psychological distress in university students. However, it is unclear which intervention is most effective. Artificial intelligence (AI) driven adaptive trials may be an efficient method to determine what works best and for whom. The primary purpose of the study is to rank the effectiveness of mindfulness, physical activity, sleep hygiene and an active control on reducing distress, using a multi-arm contextual bandit-based AI-adaptive trial method. Furthermore, the study will explore which interventions have the largest effect for students with different levels of baseline distress severity.

**Methods and analysis:** The Vibe Up study is a pragmatically-oriented, decentralised AI-adaptive group sequential randomised controlled trial (RCT) comparing the effectiveness of one of three brief, two week digital self-guided interventions (mindfulness, physical activity, or sleep hygiene) or active control (ecological momentary assessment) in reducing self-reported psychological distress in Australian university students. The adaptive trial methodology involves up to 12 sequential mini-trials that allow for the optimisation of allocation ratios. The primary outcome is change in psychological distress (DASS-21 total score) from pre-intervention to post-intervention. Secondary outcomes include change in depression, anxiety, and stress (measured by DASS-21 subscales) from pre-intervention to post-intervention. Planned contrasts will compare the four groups (i.e., the three intervention and control) using self-reported psychological distress at pre-specified time points for interim analyses. The study aims to determine the best performing intervention, as well as ranking of other interventions.

**Ethics and dissemination:** Ethical approval was sought and obtained from the UNSW Sydney Human Research Ethics Committee (HREC A, HC200466). A trial protocol adhering to the requirements of the Guideline for Good Clinical Practice [1] was prepared for and approved by the Sponsor, UNSW Sydney (Protocol number: HC200466_CTP).

**Registration details:** The trial is registered with the Australian New Zealand Clinical Trials Registry (ACTRN12621001223820).

**STRENGTHS AND LIMITATIONS OF THIS STUDY:** *Strengths:*

1. The study addresses an important clinical question using novel, advanced methods
2. The trial uses short-duration interventions designed to improve coping responses to transient stressors, which addresses the most common needs of university students
3. A value of information analysis is included to compare the value of the new trial methods with traditionalapproaches
4. Digital phenotyping is used to explore smartphone sensor information with clinical outcomes

*Weaknesses:*

1. More than 12 mini-trials might be required to determine the ranking for the interventions
2. The interventions may prove to be of the same level of effectiveness for each level of severity
3. Interventions other than those examined in this study, such as CBT, may be more effective and remain untested
4. The methodology assumes that the three digital interventions are configured to deliver similar doses and/or have approximate fidelity with standard methods

## INTRODUCTION

University students experience a disproportionate level and burden of psychological distress compared to both age-matched peers and the general adult population [2, 3]. Almost half of Australian university students report moderate or high levels of depression, anxiety or stress symptoms [4], and at least two thirds of students experience subclinical symptoms [5]. Prevalence rates are remarkably consistent across university settings [4, 6], and have remained largely unchanged for the last three decades [2]. Psychological distress is not only linked to the development of serious mental disorders, such as major depressive disorder, but is associated with early withdrawal from university study, impaired academic performance, alcohol intake, cigarette smoking, and increased risk of suicidal ideation and behaviour [2].

One approach to help target and reduce high rates of psychological distress is to deliver strategies that modify subjective responses to perceived stressors. Meta-analytic evidence confirms the potential usefulness of a range of interventions, including mindfulness-based interventions, physical activity, and sleep hygiene, to reduce anxiety and depression symptoms in university students [7–10]. However, of the wide range of interventions available for university students, it is unclear which intervention is most effective, whether interventions show differential effectiveness for mild, moderate and severe distress, and whether specific interventions are more effective for individuals with specific symptom clusters of anxiety, depression and stress. In addition, established interventions typically rely on face-to-face delivery and are often lengthy, which means they are resource intensive and difficult to deliver at scale.

Recent evidence shows that delivering interventions via smartphone apps offers a potentially feasible and scalable way to reduce psychological distress in university students, and young people more generally [10]. Being able to offer short-duration, targeted intervention in response to transient stressors, such as examinations or the transition from secondary school to university, may be particularly useful for university students. Although previous studies have included mixed delivery modes, including both face-to-face and digital self-guided interventions [8], none have compared all-digital interventions delivered using participants’ own smartphone devices.

Typically, randomised controlled trials (RCTs) are used to compare the efficacy of different interventions, or to compare the effects of an intervention with a control group. While RCTs are considered the gold-standard test of interventions and have led to a large and growing body of evidence supporting different psychological and lifestyle interventions for distressed university students, they also have some limitations. RCTs are often lengthy, expensive, and time-consuming to conduct, and are often underpowered to detect group differences, especially when comparing different active interventions. They often offer relatively little information about which individual might respond best to a specific intervention given their symptom severity, profile of symptoms, and/or demographics. These challenges underline the importance of looking for new ways to explore treatment efficacy, while preserving the rigour of a traditional RCT.

In artificial Intelligence (AI)-driven adaptive trials, instead of one large trial, we perform a series of “mini-trials” where the results of each feed into the next [11–13]. At each step, AI methods are used to a) Update an underlying model of the effectiveness of the interventions under evaluation and b) Alter the proportion of participants allocated to each intervention in the next mini-trial. Under this scheme, progressively fewer participants are allocated to less effective interventions in later mini-trials. Importantly, the sequence of mini-trials can stop as soon as the estimates of intervention effect become certain; potentially much earlier, and involving fewer individuals than a traditional RCT. In this trial, we will use contextual multi-arm bandit, which is a specific type of AI algorithm [13]. The aim of the contextual multi-arm bandit AI method is to identify the most effective intervention for a group as quickly as possible, to explore the intervention outcomes enough to ensure that one (or more) are not discarded from the trial until the best performing interventions emerge, and to perform trials to maximise statistical power while controlling false detection rates.

AI-driven adaptive trials promise to provide a quicker, and more efficient alternative to RCTs, particularly when there are multiple potentially effective options, and when it is important to determine which treatment option is best for a particular cohort of people. Compared to RCTs, adaptive trials have been argued to: 1) require fewer participants to estimate the effectiveness of an intervention [13], 2) reach a definitive conclusion earlier so that the best treatment can be offered sooner to the broader population, and 3) stop recruitment to futile interventions early, and 4) identify interactions between different interventions and different patient sub-groups [14–16]. Although first discussed over three decades ago, adaptive trials have only recently been introduced in health settings, but have recently been successfully applied in a cluster-randomised controlled trial of physical activity promotion interventions in general practice [17]. To our knowledge, AI-driven adaptive trials have never been used in the mental health context.

In the Vibe Up study, our primary aim is to use AI-driven adaptive trial methods to determine which out of three brief, two-week digital self-guided interventions (mindfulness, physical activity, or sleep hygiene) or active control (ecological momentary assessment) [18] lead to the greatest reductions in psychological distress in Australian university students. Our aim is to identify the most effective intervention within three separate ‘cohorts’ or ‘groups’: participants with normal/mild distress, moderate distress, or severe distress. If this goal is achieved, we aim to identify the second most effective intervention within each cohort. The Vibe Up study will run as a sequence of mini-trials, where participants are allocated to one of the four groups and complete self-report measures to assess how much their distress changed from pre-to post-intervention. Information about outcomes in each of the interventions (or control group) for mildly, moderately and severely distressed participants gained from one mini-trial are used to update the algorithm, which in turn will adjust how many participants are allocated to each of the four interventions in the next mini-trial. In this study, the AI algorithm has two goals: with the smallest number of participants, 1) to identify the best performing intervention within a severity group (mild, moderate, severe), and 2) to maximise the benefits for participants during the trial period.

Our second aim is to test the value of this novel trial approach in the mental health setting. We are specifically interested in establishing whether the AI-driven adaptive trial methodology is an efficient method in comparison to the traditional RCT in determining the effectiveness of the interventions. The trial will encompass an economic evaluation with a value for information analysis to determine whether the AI-adaptive trial approach yields more value compared to a traditional four-arm RCT. In theory, allocating participants using the AI-adaptive trial approach will result in fewer participants required to rank interventions by their effect size compared to a four-arm RCT strategy. This will reduce trial participant recruitment costs, although administration costs for the AI-adaptive trial approach may differ compared to an RCT. It will also variably affect the confidence intervals around each estimated effect size. The economic evaluation will therefore seek to compare the change in decision uncertainty from the AI-adaptive trial approach compared to a traditional four-arm RCT strategy.

The study also incorporates theoretically driven sub-studies focussing on assessing resilience of the students to negative affect using ecological momentary assessment (EMA) and exploring the potential for smartphone-based passive sensing of activity (digital phenotyping) to predict changes in distress symptoms. We also investigate whether the interventionshave differential effects on depression, anxiety, and stress.

The primary outcome variable is psychological distress (as measured by the DASS-21 total scores) post-intervention, relative to pre-intervention. Secondary outcomes variables include depression, anxiety, and stress (as measured by the three subscales of the DASS-21) at post-intervention, relative to pre-intervention. Other secondary variables are self-reported physical activity, sleep quality, mindfulness, and engagement with the interventions.

Based on previous meta-analytic evidence, we expect that the mindfulness and physical activity interventions will be more effective than the sleep hygiene and the active control interventions in reducing overall psychological distress in the sample (as measured by the DASS-21 total scores). We expect to identify differences in intervention efficacy according to baseline distress levels measured by the DASS-21 (mild, moderate, severe). Exploratory analyses comparing intervention effects for symptom clusters of depression, anxiety, and stress will be conducted. We expect that the AI-driven optimisation design will reduce decision uncertainty compared to a well-designed four-arm RCT that aims to establish superiority of particular interventions and any differential effects of severity on outcomes.

## METHODS AND ANALYSIS

### Trial design

This is a sequential randomised controlled trial using bandit-based response adaptive allocation to compare (on an intention-to-treat basis) the effectiveness of three brief, two-week digital self-guided interventions and an active control group in reducing self-reported psychological distress in Australian university students. The group sequential design will be executed as a sequence of up to twelve, ‘mini-trials’ each recruiting a sample of at least 120 participants. There will be no pre-defined upper limit for recruitment into each mini-trial. Each participant will be eligible to take part in one mini-trial only, with each trial lasting four weeks. Once participants have screened eligible for the study, they will complete a baseline assessment on their smartphone, two weeks of daily EMA, and a second assessment at two weeks post-baseline. Participants who do not complete the second assessment will not proceed to the intervention period of the trial. Next, they are allocated by the AI algorithm to one of the three intervention groups, or the EMA control which they receive for two weeks. Finally, all participants complete a post-intervention assessment at the end of the four-week period.

### Patient and Public Involvement Statement

The study was conceived and designed by a multidisciplinary research team consisting of clinical psychologists, software engineers, computer scientists and user experience experts. Research questions and outcome measures were derived in consultation with the target population (university students) through one-on-one consultations. Thirty-three university students experiencing psychological distress were involved in this process. They provided feedback on the initial designs of the smartphone app and the appropriateness of the intervention content and language. Individual participants will receive a summary of their wellbeing status at the end of their participation in the study. Final study results will be aggregated and presented in published manuscripts and national conference presentations. Results will also be published on the study website.

### Participants

The trial aims to enrol a total sample of approximately 1200 adult university students with mild to severe psychological distress according to the Kessler-10 at recruitment but without psychotic spectrum disorders or significant suicidality.

### Inclusion criteria

Participants must satisfy the following criteria at screening:

- Adults aged 18 or older;
- Currently residing in Australia and planning to be resident throughout their study period;
- Currently enrolled at a higher education institution;
- Advanced, fluent or native English speaker;
- Own an eligible personal smartphone (iPhone 6S/Android 5 or later) with active mobile number and internet access;
- Self-rated psychological distress on the Kessler Psychological Distress Scale, 10-item version [19] (K-10) scoring *≥* 20 [20], indicative–of “likely to have a mild (or more serious) mental disorder” [21]–at screening. The K-10 is a widely-used, validated tool for assessing psychological distress in adult populations and K-10 scores are strongly correlated with mental illness cases in community samples [22].

### Exclusion criteria

Participants will be excluded at screening if they:

- Indicate high levels of self-rated suicidal ideation (scoring *≥* 21, “high ideation”) on the Suicide Ideation Attributes Scale [23], a reliable and valid measure for assessing suicidality in general adult populations;
- Report a current active diagnosis of psychosis or bipolar disorder;
- Have already completed a previous mini-trial;
- Indicate major disruptions or events in the next 2 months which may make it difficult to take part in the study;
- Indicate plans to travel outside of Australia in the next 2 months;
- Indicate that they would be unable to safely undertake a physical activity intervention if allocated to receive this treatment.

Participants will not be restricted from receiving any other treatments during the trial but will be discouraged from undertaking new psychological therapies during the four-week trial period.

### Recruitment

The trial will be promoted via targeted paid social media advertisements placed on Facebook and Instagram; platforms identified as effective for research recruitment in young adults [24, 25]. Advertisement text was developed through formative interviews with university students. Interviews identified two themes to which potential participants might be receptive: improving personal resilience to university-related stressors, and the opportunity to contribute to improved community mental health through digital innovation. Advertisements will be targeted by age and keywords/community interests indicating affiliation to higher education institutions and will link potential participants to a study website containing study information materials and directions on how to complete online self-screening and consent. Reflecting investigator experience with similar studies, the trial will allow up to $4.50 USD to be spent on advertising per eligible enrolled participant.

Because the group-sequential design requires a stream of potential participants, performance of the advertising strategy will be monitored. Review will focus on two goals: achieving a consistent sample size per mini-trial; and trying to ensure balanced representation by presenting level of psychological distress and by gender (recognising the risk of underrepresentation of male-identifying participants in youth mental health trials [26]). Descriptive statistics concerning screening and consent rates in response to different targeting criteria and combinations of advertising text and imagery will be collected and reviewed. Responses to a single-item screening questionnaire indicating where participants heard about the study will also be assessed. Recognising the risk in a group-sequential design that adjustments to recruitment strategy introduce group-specific biases, we will aim to improve (in a prospective fashion) balanced representation by gender/distress severity rather than trying, for example, to redress a deficit in the overall sample by focussing on recruiting primarily males, or those with very high distress, for a particular mini-trial. As males tend to be more difficult to recruit into studies [27], the adopted strategy separated recruitment of male and female participants into two different campaigns and allocated more budget to social media advertisement aimed at males.

To maximise participation in the study, we also plan to advertise via university organisations, university societies, university staff contacts, and releases through traditional media outlets.

### Interventions

The trial will evaluate three brief, self-directed interventions delivered via a software application (app) installed on participants’ personal smartphones (see Table 1 for details). Each intervention is entirely separate but designed to be loosely matched on ‘dose’ and required effort over a 14-day period. Each intervention consists of brief modular information covering key concepts, delivered as infographics, structured activities (e.g., practising mindfulness with guided audio), and a ‘frequently asked questions’ section including tips, safety advice and answers to common questions associated with that intervention. In addition, an in-app support page is available with contact details of mental health support for participants. Participants are also asked to complete a daily log relevant to each intervention: for mindfulness, time spent practising mindfulness; for physical activity, time spent being physically active; for sleep, hours slept the night prior; and for EMA (active control), current affect experience including experience of both positive and negative emotions. Each participant will be allocated to receive one intervention, once. Evidence supporting the selection of the chosen interventions is included as Appendix A. All interventions are unlocked and available to participants at the end of each mini-trial.

**Table 1.**
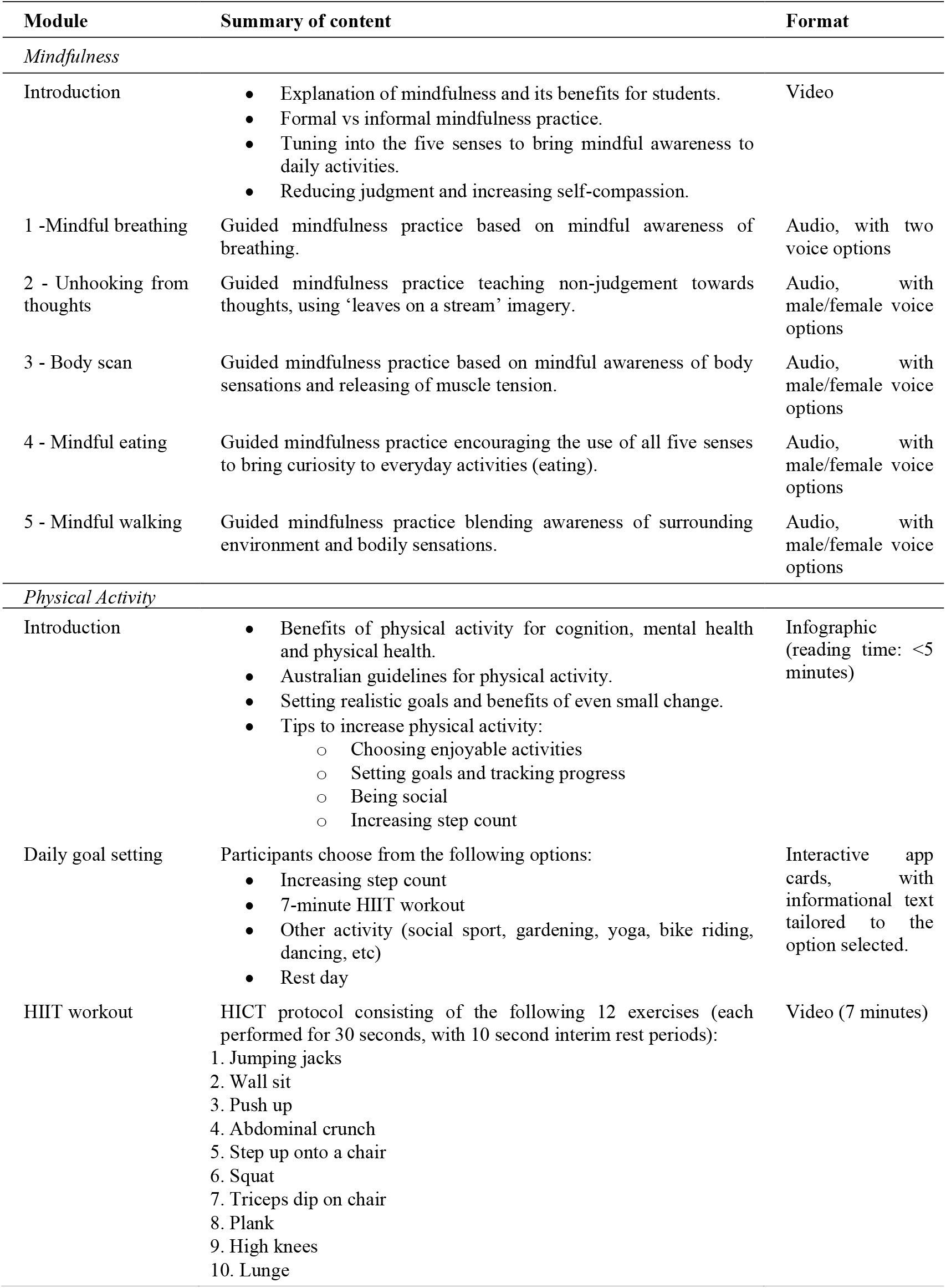
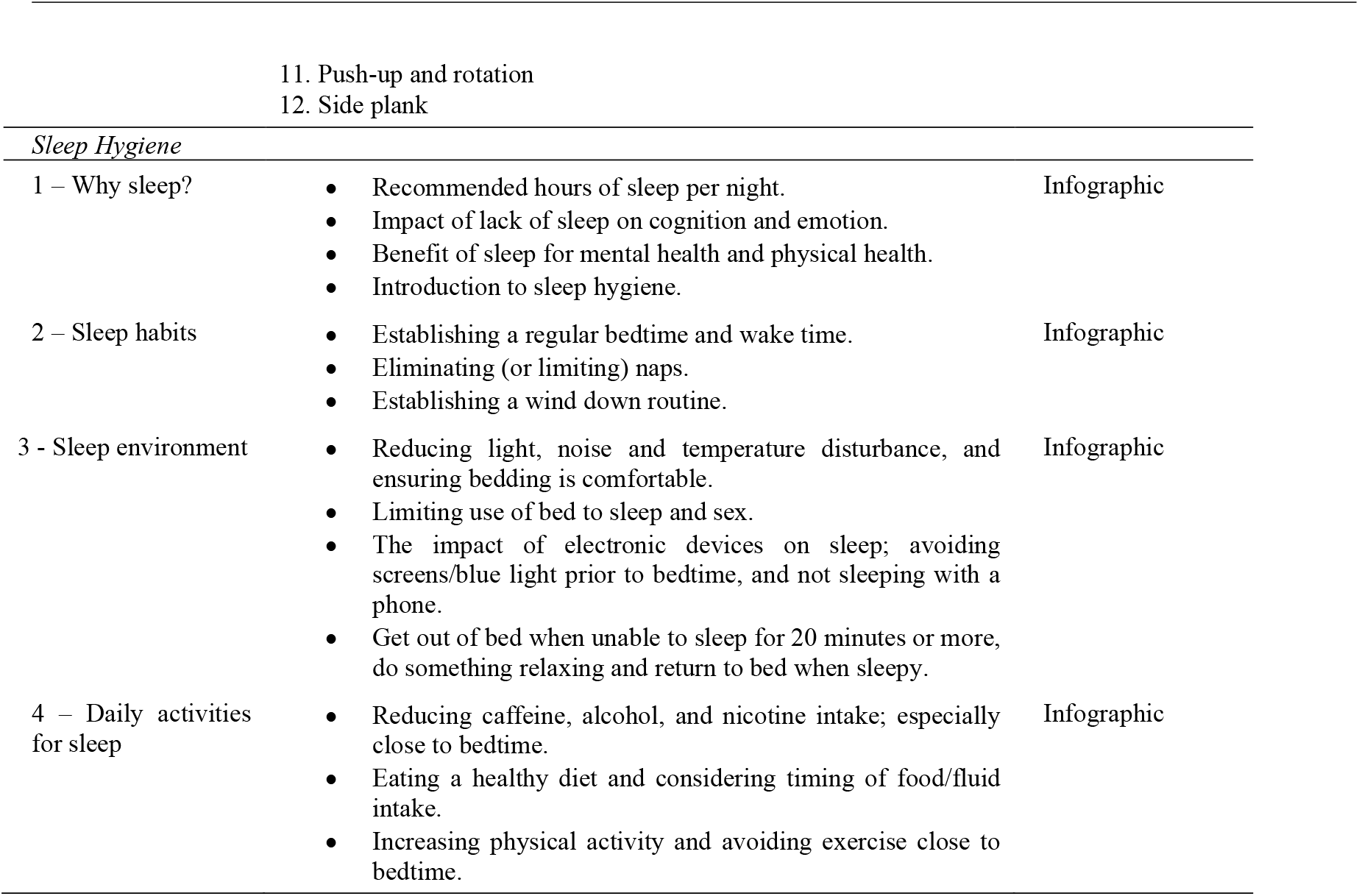
Interventions

#### Mindfulness intervention

Mindfulness is the “awareness that emerges through paying attention on purpose, in the present moment, and non-judgementally to the unfolding of experience moment by moment [28].” Mindfulness meditation can be instructor-facilitated or self-guided (e.g. using a course of guided audio meditations to learn mindfulness techniques [29]). The mechanisms of mindfulness meditation are distinct from relaxation training [30] and include changes in attention, emotion regulation, sensory awareness and self-awareness [31].

The trial mindfulness intervention consists of an introductory video that includes instructions for bringing mindful awareness to daily activities, followed by a set of five 3-5 minute instructor-guided audio meditations that focus on mindful awareness of breathing; noticing, awareness and acceptance of thoughts (‘leaves on a stream’ exercise); attending to bodily sensations (body scan); and mindful eating and walking. Encouragement to adopt a non-judgemental, accepting, and self-compassionate response to present moment awareness is weaved throughout the practices. Participants are given the choice of a male or female narrator, both of whom are Clinical Psychologists experienced in delivering mindfulness interventions. Audio recordings are released in a structured sequence with a one-day gap between each. Once released, participants can access audio and video recordings as much as they wish.

#### Physical activity intervention

The Vibe Up Physical Activity intervention starts with an introductory infographic that outlines the benefits of physical activity, Australian guidelines for physical activity, setting realistic goals and incremental change, and practical suggestions for increasing daily physical activity. The intervention was designed in consultation with Exercise Physiologists who specialise in mental health. The Vibe Up Physical Activity intervention prompts participants to choose a goal each day to increase their physical activity. An evidence-based seven-minute high-intensity interval training (HIIT) protocol is provided as one option for increasing physical activity [32]. The trial intervention delivers this as an instructor-led video of a qualified Exercise Physiologist presenting a series of exercises. Participants are asked to complete some form of physical activity on most days during the two-week intervention period. The intervention also incorporates infographic-based psychoeducation about the benefits of exercise, how exercise can be integrated into everyday life, and anchoring information about expected levels of exercise intensity during the programme. This is presented for review as a structured ‘onboarding’ process at the start of the intervention.

#### Sleep hygiene intervention

Vibe Up Sleep Hygiene is a brief, self-guided sleep hygiene intervention including basic elements of stimulus control. Sleep hygiene refers to the set of daily living activities that are necessary to maintain good quality sleep and full daytime alertness [33]. Although there are recognised common determinants of poor sleep relating to arousal (e.g. caffeine ingestion) and sleep organisation (e.g. excessive bedtime variation), the activities that influence sleep either positively or negatively substantially vary from individual to individual [33]. The purpose of sleep hygiene education (SHE) is to help individuals identify the specific behaviours and habits that promote their own sleep and implement these, while eliminating/reducing those that disturb sleep [34]. Stimulus control seeks to reduce, for those with sleep problems, anxiety or conditioned arousal associated with going to bed [35].

The trial intervention is a programme of four infographic-based information modules addressing, respectively: the importance of sleep; positive sleep habit formation; creating a sleep-promoting environment and how daily activities and diet affect sleep. Modules are released in a structured sequence with a two-day gap between each. As each module is released, participants are prompted to review its contents and identify practical ways to apply it to their sleep hygiene practices. Once released, participants can access module content as frequently as they wish.

#### Mood tracker active control

The Vibe Up Mood Tracker uses ecological momentary assessment (EMA) to characterise and quantify the individual profiles of dynamic affect experience amongst university students with elevated psychological distress. Mood trackers have often been used in conjunction with psychological therapies, including mindfulness-based interventions [36], to help monitor participants’ progress in the treatment. It captures individuals’ subjective experience of emotions to inform the changes in the state of mind and brain function. Previous research indicates that specific rhythms of affect, including ongoing mood instability and persistent negative affect, are strongly associated with the onset and progress of mental health issues such as depression, anxiety disorders [37, 38]. However, the dynamic change of emotions and its corresponding responses are often neglected in cross-sectional measurements. The thrive in mobile technologies provides a new avenue to detect the affect rhythm via real-time self-assessment, known as ecological momentary assessment [18]. Although it is still debatable whether repeated self-assessment itself may alter individuals’ affect experience [39], it is generally acknowledged that self-monitoring has minimal impact on health symptoms and mostly in the short term [40, 41], thus being used as the active control in the current study.

The Vibe Up Mood Tracker runs on a blended EMA protocol consisting of signal contingent and event-contingent EMA. For the signal-contingent EMA: Two daily random prompts will be generated by the study app at a random time within two windows: morning (08:00-10:00) and evening (19:00-21:00) according to the participants’ local time. The participants will have up to 60 minutes to respond to this prompt, with a reminder sent after 30 minutes to those not having responded to the initial prompt. For the event-contingent EMA: The participants will be able to log EMA measurements at any time (e.g. in response to self-identified exposures to negative stressors). If a participant initiates an event-contingent recording within the 08:00-10:00 or 19:00-21:00 windows, then no signal contingent prompt will be generated within that window, regardless of whether they complete their self-initiated EMA response. Each EMA prompt contains questions regarding the current feelings (positive affect and negative affect) and likelihood of responding to the selected affect(s). Recognising that participants may disclose risk information in their response, there is an annotation in the app noting that “We don’t actively monitor responses to this question, but help is always available if you need it.” plus a link to support options.

### Strategies to promote adherence

A formative user-centred design process was undertaken to identify potential strategies for promoting and sustaining engagement with study interventions and tasks. This informed the creation of a simple game-like mechanism, based on the evolution of a virtual character (Sprout), who slowly progresses from infancy to adulthood each day of the mini-trial. Embedded within the app, simple animations representing this evolution are used to provide a sense of delight and reward in response to engaging with the app (see Figure 1 for examples). In addition, a limited set of text message and email reminders are used at points in the study where there are time sensitive or mandatory tasks such as installing the study app, completing the questionnaires, accessing the interventions after allocation, and deadlines to complete these tasks.

Participants who complete the post questionnaire battery will be offered a $20 USD electronic gift token and will receive a written personalised summary report of the measurements taken during the study. Participants who additionally complete the follow-up questionnaires (at 8-weeks post intervention period) will be given the opportunity to enter a draw to receive one of three $35 USD electronic gift tokens. The amount of electronic gift tokens is not subjective to the EMA compliance rate in the current study.

### Outcome Measures

#### Primary outcome measure

##### Depression, Anxiety and Stress Scale, 21-item version

The primary outcome measure is self-rated psychological distress according to the total score on the Depression, Anxiety and Stress Scale, 21-item version (DASS-21). The DASS-21 is a reliable, valid psychometric instrument for the assessment of psychological symptoms via self-administration in non-clinical populations [42]. The DASS-21 asks participants to indicate the extent to which each symptom was experienced over the past week, using a 4-item rating scale ranging from 0 (‘Did not apply to me at all’) to 3 (‘Applied to me very much, or most of the time’). Although consisting of three subscales, addressing depressive, anxiety and stress symptoms, their suitability as a single-factor distress measure combining all subscale items has recently been demonstrated in an adolescent population [43]. Higher scores indicate higher overalldistress levels.

#### Secondary outcome measures

##### Physical Activity Vital Sign

The Physical Activity Vital Sign (PAVS) is a two-item clinical screening instrument for assessing the total time engaged in moderate to strenuous exercise over the past week in adults [44]. The instrument has been demonstrated to be valid and suitable for rapid assessment of exercise behaviour [45]. The questionnaire assesses the number of days in the past week on which moderate-to-vigorous exercise was undertaken, and the average number of minutes engaged in physical activity. This allows the calculation of the total minutes engaged in moderate to vigorous physical activity per week, and whether the participant meets the World Health Organisation Guidelines of greater than 150 minutes per week.

##### Modified Pittsburgh Sleep Quality Index Item 6

The Pittsburgh Sleep Quality Index (PSQI) is a reliable, valid 19-item instrument for assessing sleep quality over the previous month in clinical and research populations [46]. To manage participant burden, we will use only PSQI item 6 (‘During the past week, how would you rate your sleep quality overall?’) modifying this to focus on the previous week only (to be consistent with the DASS-21 and PAVS time horizons.) PSQI item 6 uses a 4-level Likert scale scoring from 0 (‘Very bad’) to 3 (‘Very good’).

##### Bespoke mindfulness measure

The mindfulness literature lacks consensus about optimal outcome measures, and measures are often lengthy [47]. Therefore, we used a bespoke single-item question to measure mindfulness. Participants were asked ‘Mindfulness is a practice where you intentionally focus your attention on what you are experiencing in the present moment, with an attitude of openness and non-judgment. During the past week, how mindful have you been?’. The question’s wording was derived from reviews of existing questionnaires [48, 49]. Participants will be asked to indicate their response using a 5-level Likert scale from 0 (‘Not at all mindful’) to 4 (‘Extremely mindful’).

##### The International Positive and Negative Affect Schedule, Short Form

The international short form (I-PANAS-SF) [50] is a 10-item scale assessing participants’ subjective experience of positive (e.g., inspired), and negative emotional states (e.g., upset, afraid). The items were rated on a 5-level Likert scale from 1 (‘Very slightly or not at all’) to 5 (‘Extremely’). Subscale scores are determined as the sum of item scores, with higher scores indicating higher levels of positive or negative affect. Reflecting on the trial focus on distress, the I-PANAS-SF is extended with two distress-focused items (feeling hopeless or calm) from the K10 to depict the momentary levels of psychological distress. To minimise burden, participants will be asked to choose which of the twelve feelings applied to them at the moment, and then rate the intensity of each of the feelings they selected. Non-selected feelings will be coded as 1 (‘Very slightly or not at all’). If a participant selected one or more feeling(s), they will be asked further bespoke questions on their likelihood to do something because of their feelings, using a 4-level Likert from 0 (‘Highly unlikely’) to 3 (‘Highly likely’.) If a participant responds 2 (‘Likely’) or 3 (‘Highly likely’), a second question will ask them to describe using free text what is that they are likely to do.

#### Additional measures

Additional measures (as seen in Table 2) will be collected either during screening, pre- or post-intervention and used as predictors in models, as mediators of intervention effect and in exploratory analyses. These will assess contextual and perception-related factors that may influence subjective distress or intervention response, such as the availability of social support, socioeconomic status and substance use. They also include measures of subjective attitudes concerning the likely success of the intervention, readiness for change, post-hoc perceptions of the interventions and technology experience/barriers to use.

**Table 2.**
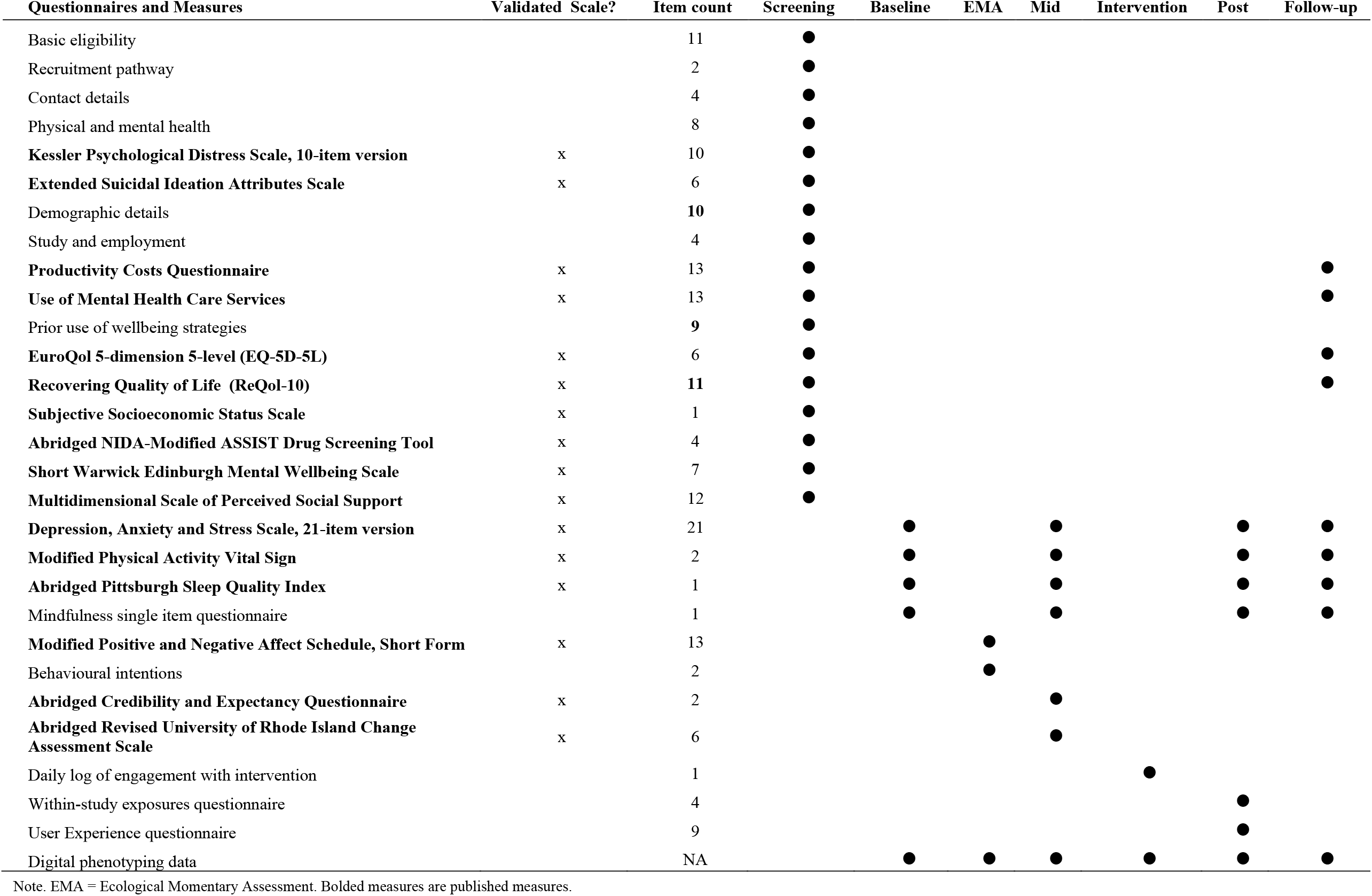
Questionnaire measures and administration

| Questionnaires and Measures | Validated Scale? | Item count | Screening | Baseline | EMA | Mid | Intervention | Post | Follow-up |
| --- | --- | --- | --- | --- | --- | --- | --- | --- | --- |
| Basic eligibility |  | 11 | ● |  |  |  |  |  |  |
| Recruitment pathway |  | 2 | ● |  |  |  |  |  |  |
| Contact details |  | 4 | ● |  |  |  |  |  |  |
| Physical and mental health |  | 8 | ● |  |  |  |  |  |  |
| **Kessler Psychological Distress Scale, 10-item version** | x | 10 | ● |  |  |  |  |  |  |
| **Extended Suicidal Ideation Attributes Scale** | x | 6 | ● |  |  |  |  |  |  |
| Demographic details |  | 10 | ● |  |  |  |  |  |  |
| Study and employment |  | 4 | ● |  |  |  |  |  |  |
| **Productivity Costs Questionnaire** | x | 13 | ● |  |  |  |  |  | ● |
| **Use of Mental Health Care Services** | x | 13 | ● |  |  |  |  |  | ● |
| Prior use of wellbeing strategies |  | 9 | ● |  |  |  |  |  |  |
| **EuroQol 5-dimension 5-level (EQ-5D-5L)** | x | 6 | ● |  |  |  |  |  | ● |
| **Recovering Quality of Life (ReQoL-10)** | x | 11 | ● |  |  |  |  |  | ● |
| **Subjective Socioeconomic Status Scale** | x | 1 | ● |  |  |  |  |  |  |
| **Abridged NIDA-Modified ASSIST Drug Screening Tool** | x | 4 | ● |  |  |  |  |  |  |
| **Short Warwick Edinburgh Mental Wellbeing Scale** | x | 7 | ● |  |  |  |  |  |  |
| **Multidimensional Scale of Perceived Social Support** | x | 12 | ● |  |  |  |  |  |  |
| **Depression, Anxiety and Stress Scale, 21-item version** | x | 21 |  | ● |  | ● |  | ● | ● |
| **Modified Physical Activity Vital Sign** | x | 2 |  | ● |  | ● |  | ● | ● |
| **Abridged Pittsburgh Sleep Quality Index** | x | 1 |  | ● |  | ● |  | ● | ● |
| Mindfulness single item questionnaire |  | 1 |  | ● |  | ● |  | ● | ● |
| **Modified Positive and Negative Affect Schedule, Short Form** | x | 13 |  |  | ● |  |  |  |  |
| Behavioural intentions |  | 2 |  |  | ● |  |  |  |  |
| **Abridged Credibility and Expectancy Questionnaire** | x | 2 |  |  |  | ● |  |  |  |
| **Abridged Revised University of Rhode Island Change Assessment Scale** | x | 6 |  |  |  | ● |  |  |  |
| Daily log of engagement with intervention |  | 1 |  |  |  |  | ● |  |  |
| Within-study exposures questionnaire |  | 4 |  |  |  |  |  | ● |  |
| User Experience questionnaire |  | 9 |  |  |  |  |  | ● |  |
| Digital phenotyping data |  | NA |  | ● | ● | ● | ● | ● | ● |

Note. EMA = Ecological Momentary Assessment. Bolded measures are published measures.

##### Short Warwick Edinburgh Mental Wellbeing Scale

The seven-item instrument (SWEMWBS) is an abbreviated version of the Warwick Edinburgh Mental Wellbeing Scale, originally designed to assess general mental wellbeing in adult populations [51]. Although SWEMWBS provides less coverage of hedonic well-being and affect, it is sensitive to psychological wellbeing, has robust measurement properties and is explicitly recommended for general population monitoring [52]. Participants are asked to rate a series of statements concerning experiences and attitudes over the past two weeks (e.g. ‘I’ve been feeling optimistic about the future’) using a 5-level Likert scale ranging from 1 (‘None of the time’) to 5 (‘All of the time’). A total score is derived by summing item scores and transforming using a published lookup table [52] with higher scores indicating higher positive mental wellbeing.

##### Multidimensional Scale of Perceived Social Support

The Multidimensional Scale of Perceived Social Support is a 12-item reliable instrument with moderate construct validity which asks participants to rate twelve statements concerning support available from a three factor structure of friends, family and significant others (e.g. ‘There is a special person who is around when I am in need.’ [53]). Participants are asked to indicate agreement with each statement using a 7-level Likert from 1 (‘Very strongly disagree’) to 7 (‘Very strongly agree’). A total score is generated as the arithmetic mean of item scores, with a higher score indicating greater levels of perceived support. Subscale scores can be generated for each of the three factors but will not be used in this trial.

##### Subjective Socioeconomic Status Scale

The Subjective Socioeconomic Status Scale is a simple self-anchoring scale that uses a visual metaphor of status – an image of a ten-rung ladder – and asks participants to locate their perceived position on the rungs. The top of the ladder is explained as representing those who ‘are the best off’, having ‘the most money, the most education and the most respected jobs’ while the bottom represents the opposite. The scale yields a score from 1-10 inclusive where 10 represents higher perceived socioeconomic status. Subjective socioeconomic status is better correlated than objective measures (such as income) with psychological variables such stress, negative affect and coping [54].

##### NIDA Quick Screen drug screening tool

The US National Institute on Drug Abuse (NIDA) Quick Screen is a four-item instrument [55] adapted from a single question screening tool for drug use in primary care [56]. The original instrument asks for the number of times that any drug has been used in the past year, while the NIDA tool asks the question separately for binge use of alcohol, any use of tobacco products, prescription drugs being used for non-medical reasons and any use of illegal drugs. Binge use is defined as five or more drinks in one day for males or four or more for females. Participants are asked to respond using a 5-item scale from: ‘Never’, ‘Once or twice’, ‘Monthly’, ‘Weekly’ or ‘Daily or Almost Daily’.

##### Credibility and Expectancy Questionnaire Items 1 and

1 The Credibility and Expectancy Questionnaire (CEQ) is a reliable six-item scale assessing two cognitive factors concerning belief in an intervention (credibility) and expectation of benefit from its use (expectancy) [57]. Expectancy, appears to be associated with observed outcomes in intervention research [57, 58]. To manage participant burden, we will use a single CEQ item to assess each of the factors. Item 1 loads on credibility (‘How logical does the therapy offered to you seem?’) and is rated with a 9-level Likert from 1 (‘Not at all logical’) to 9 (‘Very logical’). Item 6 assesses expectancy (‘How much improvement in your symptoms do you really feel will occur?’) and is rated using an 11-level numeric score from 0-100% in 10% increments. Each factor will be evaluated separately.

##### Revised University of Rhode Island Change Assessment Scale Items 19, 25-26, 29 and 30

The Revised University of Rhode Island Change Assessment (URICA) scale is a 32-item scale originally intended to assess readiness for change during psychotherapy [59]. The trial will assess three factors identified in the original scale that are relevant before starting a new treatment: seeking assistance, ambivalence towards change and taking action, selecting the two URICA items for each factor that accounted for the highest proportion of variance explained in the original study (Items 19 and 24 for seeking assistance; Items 26 and 29 for ambivalence towards change; and Items 25 and 30 for taking action). Each item is a statement relating to change readiness in the current moment (e.g. ‘I wish I had more ideas on how to solve my problems.’) and is scored using a 5-level Likert from 1 (‘Strongly disagree’) to 5 (‘Strongly agree’). Items 26 and 29 are reverse scored. A total score is generated by summing the scores with higher scores indicating greater change readiness.

#### Health economic evaluation measures

Additional measures will be collected for an economic evaluation using a health system and societal perspective. This will include measuring health outcomes and healthcare costs, along with changes in resource use outside the healthcare system. This trial will include an evaluation of productivity changes to capture whether improvements in psychological distress due to the interventions impacted workforce participation.

##### EQ-5D-5L

The EuroQol 5-dimension 5-level (EQ-5D-5L) is a generic preference-based health related quality of life tool used to estimate quality of life and to undertake cost utility analysis [60–62]. It has five descriptive dimensions, including mobility, self-care, usual activities, pain/discomfort and anxiety/depression [63]. Each dimension has five levels, including no problems, slight problems, moderate problems, severe problems and extreme problems. Responses to the EQ-5D-5L will be captured at screening and 8-week follow-up, and converted into utilities using an algorithm derived from the Australian general population [64]. Quality-adjusted life years (QALYs) for each participant will be estimated by using estimates of utilities and the area-under-the-curve method [65].

##### ReQol-10

Recovering Quality of Life (ReQoL) 10 [66] is a preference-based health related quality of life tool used to estimate quality of life for people with mental health conditions. It has been developed to be more sensitive to generic preference-based health related quality of life tools when measuring differences in mental health outcomes. It contains 10 mental health items and one physical health item. Responses to ReQoL-10 are captured at screening and 8-weekfollow-up, and will be converted into utilities using an algorithm derived from the UK general population. Quality-adjusted life years (QALYs) for each participant will be estimated by using estimates of utilities and the area-under-the-curve method.

##### iMTA Productivity Costs Questionnaire

The iMTA Productivity Cost Questionnaire [67] is a self-completed questionnaire that measures health related changes in productivity. Productivity losses related to mental ill health will be measured across three domains, including absenteeism, presenteeism and unpaid work.

##### Use of Mental Health Care Services Questionnaire

The Use of Mental Health Care Services questionnaire was developed specifically for this study to collect information from trial participants on their use of services before and after the interventions. It surveys participants to collect information on the number of services used in the last four weeks across five domains, including hospital services, out-of-hospital services, online self-help services, community based services, and medicines.

#### Additional bespoke questionnaires

There are six additional study-specific questionnaires. These are provided in Supplemental File 1 and are described below. A demographics questionnaire will solicit information about gender, sex recorded at birth, sexual orientation, Aboriginal and Torres Strait Islander status, ethnic origin, and language most used at home. A study and employment questionnaire will ask at screening about international student status; academic performance (reported as current Weighted Average Mark or Grade Point Average) and employment in a paid job concurrently with studies. During the two-week intervention period, participants allocated to an active treatment will be asked to complete an intervention-specific questionnaire asking how many minutes of, respectively, mindfulness, physical activity and sleep they achieved in the previous day. In addition, three items are included to assess whether participants had prior experience in using wellbeing strategies.

Post-intervention, a within-study exposures questionnaire will ask participants to indicate possible disruptions over the past two weeks that may have affected their ability to complete or derive benefit from the intervention in four domains: life or routine disruption; negative impacts on mental health; positive impacts on mental health; problems using the study app. Participants will be asked to rate the extent to which they agree with each of these domains expressed as a statement (e.g. ‘In the past two weeks, my life or routine was disrupted for some reason.’) using a 5-level Likert from 1 (‘Strongly disagree’) to 5 (‘Strongly agree’).

Finally, a user experience questionnaire, based on the System Usability Scale [68] and the mHealth App Usability Questionnaire [69], will ask all participants about subjective perceptions on ease of use, usefulness, satisfaction and technology problems using the study app. For those allocated to an intervention, they will additionally be asked about trust and, separately, novelty of the informational content of the intervention, the extent to which they implemented intervention suggestions in the past two weeks and their intentions to do so in the future. As for the within-study exposures questionnaire, all items are expressed as statements (e.g. ‘I found the app easy to use.’) and rated using the same 5-level Likert agreement scale.

#### Digital phenotyping

Passive sensor data will be sampled and collected by the app according to either a predetermined frequency or change in user activity (e.g., change in location, or walking to running). The passive sensor data collection is contingent on the user granting permissions on both registration and first launch of the app. These permissions may be granted or revoked by the user at their discretion through the course of the study. This data entails information about physical movements (dynamic state) of smartphones in space and includes specific data generated from inertial sensors (accelerometry and gyroscope) and from GPS sensors that determine travelled distance, geographical location, user activity (e.g. walking, running, driving, as determined by the device Operating System), and step count. These data points can be used to explore associations with changes in anxiety, depression, and stress levels within individuals, and to predict DASS-21 at endpoint.

Participants can decline all phenotyping during the trial registration process, meaning the app will not record any such data during the trial.

### Study procedure

We are planning to conduct up to 12 mini-trials. Recruitment into each mini-trial will open one week prior to the planned start date. Applicants to the study will read the electronic online information sheet and consent form, provide consent electronically during completion of the screening questionnaires on the Qualtrics survey platform. All eligible participants will be invited to install the study app via text message.

On installing the study app and registering via Time-based One-Time Password (TOTP), participants will be prompted to complete baseline questionnaires and start daily ecological momentary assessment. All subsequent study procedures will be directed via the app. Ten days later, participants will be invited to complete the pre-intervention questionnaire battery and upon completion, will be automatically allocated to one of the three interventions or the active control condition. App-generated prompts will guide participants on how to commence and subsequently undertake the interventions.

The intervention period will last two weeks. Outcomes will be measured immediately post-intervention and again at an eight-week follow-up. After completion of the post-intervention measures at four weeks, estimates of the intervention effect will be used to update the multi-arm bandit algorithm in time to perform allocation for the following mini-trial. After the post-intervention assessments, all study interventions will become available for participants to use for a maximum period of eight weeks after the trial finishes.

The trial will continue until a significant difference can be ascertained between the AI algorithm-estimated effect sizes of the most effective, and the second most effective interventions (after appropriate adjustment for repeated comparisons) within each severity cohort (mild, moderate, severe distress). If it is not possible to separate our intervention effects in this time, the trial will conclude when twelve mini-trials have been conducted. The rationale for determining the significant difference between the most effective intervention compared to the rest is clear. The rationale for establishing which is the second-best intervention is based on our view that this additional information about effectiveness will be helpful clinically and theoretically. First, it offers a second line of treatment if a person is unable to undertake the first (e.g., Unable to undertake physical activity) and it provides the opportunity to examine contextual factors that might impact on the two treatments differentially. In typical non-adaptive trials, a series of planned or post hoc analyses are frequently used to compare mean differences between different intervention types and compared to the control group. Because we seek to detect a significant difference between the best intervention and the others, and the second best intervention and those remaining, the number of mini-trials initiated will be determined by the performance of the AI algorithm in learning the differences in intervention effect between each intervention.

### Randomisation/Blinding

Computerised allocation will be performed automatically for participants who complete the baseline questionnaires. In the first mini-trial, an allocation ratio of 1:1:1:1 (mindfulness, physical activity, sleep hygiene, EMA control) will be used. Subsequent allocation ratios will be determined by the multiarm bandit algorithm. There will be no minimum per-arm allocation implying that poorly performing arms can be dropped. Participants’ group allocation will be revealed to them within the app after they complete the mid-surveys (after the EMA period).

Participants and operational staff involved in day-to-day participant management will be unblinded because the nature of the interventions mean that they cannot easily be concealed. All other investigators and trial staff will be blinded. Allocation concealment will be guaranteed by preventing access by blinded study staff to the computer system holding randomisation information; and breaking randomization codes only once primary data analysis is complete (or at the request of the Data Safety Monitoring Board). Intervention allocation codes will be generated and retained automatically by the computer system performing allocation.

### Multi-arm bandit algorithm

For the Vibe Up trial, the specified optimisation problem is to identify, with the smallest number of mini-trials/participants, the best performing intervention arm. This means that we can reject the null hypothesis that there is no difference between the best performing intervention and the other three groups in the pre-to post-intervention change scores on the DASS-21 Total. If this is successful within 12 mini-trials, the problem will be reformulated to try to establish the next best-performing intervention within the remaining trials. Assessment of whether optimisation goals have been satisfied will be made offline as part of interim analyses conducted after mini-trials 4, 8, and 12 (outlined in detail below).

The bandit algorithm used in the trial will have the following technical properties. Intervention effects will be modelled using Gaussian Process regression with zero mean function and squared exponential kernel, with baseline normalised DASS-21 score as the sole independent variable (capturing ‘context’) and within-individual pre-post DASS-21 change score as the dependent variable, treating both as continuous and real-valued. Change scores will be used here – but not for the main trial analyses – for consistency across models by ensuring that a value of 0 implies no effect (this ‘contextual bandit’ regression approach will mean that severity-contingent effects can also be estimated and compared for the interventions). After mini-trial one, which will use a fixed allocation probability of 0.25 per arm, an Upper Confidence Bound (UCB) acquisition function will be used to deterministically allocate participants to the modelled best-performing intervention given their baseline DASS-21 severity level. UCB uses a statistically rigorous scheme to balance two goals, exploitation (to maximise allocation of apparently ‘good’ intervention arms) and exploration (to collect data about all arms to improve our knowledge about their goodness).

### Sample size

The trial will recruit at least 120 participants in each of up to 12 mini-trials. To allow for attrition between screening and mini-trial commencement, recruitment for each mini-trialwill continue until at least 120 individuals have been screened eligible. Assuming that up to one third of participants do not respond to the subsequent invitation to install the study app and complete baseline questionnaires, this will yield at least 80 participants starting the mini-trial (i.e. 20 per arm, assuming the mini-trial 1 allocation ratio of 1:1:1:1). Attrition after baseline of up to 20% is allowed, resulting in expected completion of post-intervention assessments of 64 (i.e. approximately n=16 per arm).

Traditional power estimation tools are not well suited to adaptive allocation schemes. Assuming, conservatively, that the study was able to recruit a minimum of 192 participants in each of the two best-performing intervention arms over 12 mini-trials, it would have power of 80% to detect a two-tailed effect size of 0.29 (small-moderate) between the groups (with alpha set at 5%) in a combined analysis of all availabledata.

### Statistical analysis and data management Analysis of primary outcome

For analysis, participants will be categorized into one of three clinically-relevant severity groups according to their baseline DASS-21 total score (following the procedure outlined in the DASS manual): Normal/mild symptoms, Moderate symptoms, Severe/extremely severe symptoms [70]. This approach will allow exploration of whether the most effective intervention(s) differ according to severity.

Mixed model repeated measures analysis of variance [71] will be used to compare the four groups. Analysis of the primary endpoint will be based on an intention-to-treat analysis strategy, under the assumption that missing data are missing at random. Participants must have downloaded the app and completed both the baseline DASS-21 and pre-intervention DASS-21 assessments to be included in the intention-to-treat sample. An unconstrained variance–covariance matrix will model within-individual dependencies. Satterwhaite’s method [72] will be used to adjust degrees of freedom. For each group, planned contrasts will compare the difference in self-reported psychological distress from pre-to post-intervention (or control period) as measured by the DASS-21 total score in each mini-trial. Any required transformation of scores to meet distribution assumptions of analysis will be undertaken with results from transformed data forming the basis of judgements of statistical significance. Choice of transformation will bemade on review of the data from the first mini-trial and be used in all severity group and all subsequent mini-trials. A significance level of 0.05 will apply to tests conducted in the mini-trials.

### Interim analyses

We will conduct interim analyses three times during the trial period, after mini-trials 4, 8 and 12, using the full available data in the intent-to-treat sample. These analyses will determine whether a particular intervention is more effective in reducing distress from pre-to-post-intervention, relative to the other groups, within each clinical severity group (i.e., mild, moderate, severe). Once an intervention has been found to be the most effective within a severity group, it will be removed from the list of interventions available for recommendation by the optimisation algorithm for the remaining mini-trials. This allows the optimiser to focus its allocations for the rest of the mini-trials to the remaining interventions (or control group).

For example, if the interim analysis reveals that for participants with severe distress, the physical activity intervention is more effective than the other three groups (mindfulness, sleep, EMA), it will be removed from the list of possible interventions that participants with severe distress can be allocated to in the remaining mini-trials. This will enable the optimiser to determine the next best intervention for people with severe distress between the three remaining groups.

The intention of this approach is that at a future interim analysis, comparisons can be made to find the second most effective treatment within the severity group. The process is repeated until the second-best intervention is identified, and the third most effective and so on. Although attempts will be made to rank the effectiveness of the four groups from most to least effective within each severity cohort, there is no guarantee that the full ranking will be complete by the end of the 12 mini-trials.

Comparisons of multiple treatments at multiple timepoints can lead to inflation of Type 1 error (false positive) rates, which need to be corrected for. To control for Type 1 errors that arise from sequential hypothesis testing, an alpha spending function will be applied to distribute the Type 1 error across the three interim analyses. Various spending functions exist such as Pocock, O’Brien-Fleming, Demets & Lan [4]. In our study, we will conduct the first hypothesis test at about 33% of the way during the experiment period (after mini-trial 4), the second at about 66% of the way (after mini-trial 8) and the third after the final mini-trial (100% of the way through). The information fraction for the alpha spending function is the fraction of participants data so far, compared to the expected total number of participants over the experiment period. Appropriate adjustments to the critical alpha spending p-value will be made to ensure the cumulative Type 1 error is maintained at 0.05. In addition, to control the increased Type-I errors due to multiple hypothesis tests, we will apply the Benjamini-Hochberg Correction in the critical p-values values [5]. This means that the alpha spending p-value is adjusted for each of the multiple tests. For example, in the first intermittent test, there will be 4 treatments to compare for each cohort. This totals 6 comparisons (done via t-tests). As such the alpha spending p-value for this test is adjusted according to the Benjamini-Hochberg method.

At each intermittent analysis, a multiple hypothesis test is conducted (separately for each severity group) to determine whether, for the currently active interventions, any of them is significantly better at improving the DASS score than the other groups.

For a treatment to be removed from the list of available treatments for the severity group (and be deemed the most effective treatment option), it must emerge significantly better (one-sided Welch t test with Satterthwaite adjusted degrees of freedom) in pairwise comparisons between it and every other active intervention or control group, within the cohort. Significance is specified as returning a p-value less than the critical alpha spending p-value (with the Benjamini-Hochberg p-value adjustment).

### Additional analyses

Mixed effect logistic or Poisson regression models will be used to assess if baseline psychological distress and suicidal thoughts/behaviour are associated with engagement with EMA. The effect of time-varying responses to feelings, momentary affect, and changes in self-reported psychological distress and suicidal thoughts and behaviour will also be explored using mixed effects regression models. The relationship between momentary affect, psychological distress, exercise, and sleep quality at a given time interval will be assessed by mixed effect regression models. Descriptive analyses will be used to examine compliance and reactivity of the EMA.

Machine learning will be used to analyse digital phenotyping data to: 1) explore whether any novel behavioural factors predict the study primary endpoint; and 2) investigate within-individual behavioural signals that predict individual changes in self-reported distress or affect measured using EMA.

Raw data collected from sensors will first be pre-processed and feature extraction techniques applied. Parts of this data are high dimensional, such as acceleration and angular acceleration, and therefore signal processing techniques will be used to extract low-level features. The data will then be investigated both separately and in conjunction with survey responses and EMA results to identify and select an optimal set of variables for machine learning algorithms. This will allow us to develop predictive models of participants’ mental health state(s) across various stages of the study.

In terms of economic evaluation of potential benefits of running AI-adaptive trials, the expected value of this methodology is the potential reduction in the probability of making a wrong funding decision (i.e., the intervention is not the most cost-effective intervention) multiplied by the average consequence of being ‘wrong’ (i.e. how resources may be better allocated). This benefit is compared with the cost of the trial itself. If the expected benefit exceeds the expected cost, then there is a net gain to using the AI-adaptive trial.

A value of information analysis (VoI) will be used to estimate if the bandit-based adaptive trial represents better value than conducting a traditional RCT [73]. This will be conducted ex-ante to assess whether conducting the AI-adaptive trial would be more valuable compared to a RCT before it is conducted, and ex-post to assess whether the AI-adaptive trial was more valuable in terms of reducing the need for further research compared to a RCT.

The primary challenge with this economic evaluation is that a traditional RCT is not being run alongside the AI-adaptive trial, so the uncertainty reduction in the intervention rankings and differences in trial costs cannot be directly compared. For the ex-ante VoI, the mean and uncertainty (standard errors) surrounding the estimates of QALYs and costs experienced with each intervention and trial design will be estimated using reported interventions effectiveness in the published literature, combined with estimates of the sample size required to conduct an RCT to show significant differences in the treatment effects between interventions. For the AI-adaptive trial expost analysis, the mean and uncertainty surrounding the estimates of the mean QALYs and costs experienced with each intervention will be based on the results of the AI-adaptive trial. For the RCT ex-post analysis, the expected mean and uncertainty surrounding the estimates of the mean QALYs and costs that would have been experienced using a traditional RCT will be estimated using data collected in the first mini-trial.

Costs will include those associated with developing the AI-adaptive trial algorithm (for the AI-adaptive trial arm only), analysis, participant recruitment, app dissemination, app maintenance and hosting, students’ productivity students’ mental health care services use, and the time of participants using the app.

The expected value of perfect information (EVPI), expected value of sample information (EVSI), and expected net benefit of sampling (ENBS) would be estimated using analytic methods [74–76]. A willingness to pay threshold of $50,000/QALY gained will be assumed [77]. We will estimate the population size that may benefit from the app based on the inclusion criteria. The VoI will be conducted on a time horizon according to the assumed life expectancy of the intervention based on estimates of the duration of time until the app needs to be redeveloped.

If the EVPI is inferior to the potential cost ofresearch, then there is no value in conducting new research, and the VoI analysis can stop. If the EVPI is superior to the research costs, then EVSI will be estimated and compared to the trial costs to compute the ENBS for the AI-adaptive trial and the hypothetical RCT. If the ENBS from a traditional RCT is estimated to be less than the ENBS for the AI-adaptive trial, the latter will have produced a net societal gain, supporting evidence for its use [78].

### Data management

All trial data will be collected electronically using online questionnaire software and the Vibe Up app which will transfer collected data automatically to a cloud database. To avoid accruing data plan costs for participants, data will, by default, be transmitted to the research team only when each mobile device is connected to a Wi-Fi network. To ensure that no data are lost, the app will securely store collected data until it has been successfully transmitted to the server. After collection, all data will be transferred on a scheduled basis by the research team to a secure network drive for storage and backup. A combination of technical and procedural access controls, documented in a Research Data Management plan, will be used to restrict access to data and specify the purposes for which it can be used. Participant identifiable details, such as contact information, will be held separately from other trial data. Identifiable information necessary for the administration of the study and/or participant safety follow up, such as contact details and participant follow-up records, will be accessible only to named members of the research teams involved in participant administration/safety responses. Identifiable information will not be used for any study analysis. Upon completion of this project, all information will be retained for 15 years. Procedures for data archival and destruction will follow the then-current UNSW Sydney procedures and the Australian Code for the Responsible Conduct of Research [79].

### Ethics, oversight and dissemination

Ethical approval was sought and obtained from the UNSW Sydney Human Research Ethics Committee (HREC A, HC200466). The trial Sponsor is UNSW Sydney. The trial is registered with the Australian New Zealand Clinical Trials Registry (ACTRN12621001223820). Any protocol amendments will be subject to approval by the ethics committee and will be recorded in the ANZCTR registry. Annual reports of study progress will be submitted to the HREC and Sponsor.

The study design and intervention materials were developed in conjunction with those with lived experience of anxiety and depression. The study oversight includes a Stakeholder and Advisory Board, which comprises psychiatrists, clinicians, lived experience leaders, researchers, data scientists and service providers.

A participant safety response procedure was established to address potential psychological safety risks, including elevated suicidal ideation at initial screening or other unprompted disclosures (for example, to the study email account) of significant distress. Under the protocol, risk disclosures will be managed by offering a phone call with a study clinician and a range of self-referral options, such as consulting a general practitioner and links to crisis support services. Oversight of any psychological safety events, any other adverse events and progress towards trial outcomes will be provided by an independent Data Safety Monitoring Board. The board will meet regularly throughout the trial and is specifically required, if necessary, to make recommendations to the Sponsor on whether to continue, modify, or stop the trial.

Access to the full study protocol and associated written procedures is available on reasonable request. Details of the specific implementation of the bandit algorithm will be available on request once the study is complete. Access to participant-level data will be subject to the governance procedures of a planned data repository, accessible to researchers and non-commercial users, that will contain the data arising from this study. Study source code, including that of the study app and trial administration platform, are not publicly available.

Study findings will be disseminated principally via peer-reviewed publications in scientific journals and via academic conference presentations. Information materials and a dissemination programme will be developed to share learning around bandit-based response adaptive randomisation with potential clinical research users. Data used in academic outputs and training materials will be in aggregate form only. There are no funding-related restrictions on how study information can or will be disseminated.

## Data Availability

Access to participant-level data will be subject to the governance procedures of a planned data repository, accessible to researchers and non-commercial users, that will contain the data arising from this study

## Authors’ contributions

SV, KH and HC conceived the study and secured funding with assistance from JH, JB, AWS, JN, RV, KM, SG, SR, TT, TQ and HC. KH led development of the study protocol and wrote the first draft of this manuscript. ES, WYZ, JH, JN, LH, RL, TQ and SG played key roles in the design and development of study processes. ES developed the intervention designs and content with support from MB, JB, JH, SR and JN. JH devised the ecological momentary assessment study component. LH led development of the technical platform, with support from RL, SB, RV and KM. SG and MS led development and implementation of the bandit algorithm, with support from SR, SV and AMcK. HC specified the economic analyses. All authors provided critical input throughout the development of the protocol. All authors reviewed the manuscript and provided comments. All authors approved the manuscript prior to publication.

## ACKNOWLEDGEMENTS

We would like to thank Adryon Joubert-de Villiers, Jenny Vuu, Caroline Fitzgerald, Matthew Slarke, Marya Bautista, Vera Kravchuk, Matt Lee, Dean Winder and Sandrine Chevassu for their significant contributions to the design and development of the study app and digital intervention content. We are also very grateful to Debbie Agnew and Zoe Jenkins for their advice on social media recruitment strategy and content, and to David Jung for helping secure data governance approvals. We would also like to thank the university student organisations and counselling services for their feedback on design and intervention content.

## REFERENCES

1. International Council for Harmonisation of Technical Requirements for Pharmaceuticals for Human Use (ICH), Integrated Addendum to ICH E6(R1): Guideline for Good Clinical Practice. 2016.

2. Sharp, J. and S. Theiler, A Review of Psychological Distress Among University Students: Pervasiveness, Implications and Potential Points of Intervention. Int J Adv Couns, 2018. 40(3): p. 193–212.

3. Cvetkovski, S., N.J. Reavley, and A.F. Jorm, The prevalence and correlates of psychological distress in Australian tertiary students compared to their community peers. Aust N Z J Psychiatry, 2012. 46(5): p. 457–467.

4. Larcombe, W., et al., Prevalence and socio-demographic correlates of psychological distress among students at an Australian university. Studies in Higher Education, 2016. 41(6): p. 1074–1091.

5. Stallman, H.M., Psychological distress in university students: A comparison with general population data. Aust Psychol, 2010. 45(4): p. 249–257.

6. Adlaf, E.M., et al., The Prevalence of Elevated Psychological Distress Among Canadian Undergraduates: Findings from the 1998 Canadian Campus Survey. J Am Coll Health, 2001. 50(2): p. 67–72.

7. Huang, J., et al., Interventions for common mental health problems among university and college students: A systematic review and meta-analysis of randomized controlled trials. J Psychiatr Res, 2018. 107: p. 1–10.

8. Barnett, P., et al., The efficacy of psychological interventions for the prevention and treatment of mental health disorders in university students: A systematic review and meta-analysis. Journal of Affective Disorders, 2021. 280: p. 381–406.

9. Young, C.L., et al., Efficacy of online lifestyle interventions targeting lifestyle behaviour change in depressed populations: A systematic review. 2018, Sage Publications: US. p. 834–846.

10. Parker, A.G., et al., The effectiveness of simple psychological and physical activity interventions for high prevalence mental health problems in young people: A factorial randomised controlled trial. Journal of Affective Disorders, 2016. 196: p. 200–209.

11. Allender, S., et al., Bayesian strategy selection identifies optimal solutions to complex problems using an example from GP prescribing. npj Digital Medicine, 2020. 3(1): p. 7.

12. Greenhill, S., et al., Bayesian Optimization for Adaptive Experimental Design: A Review. IEEE Access, 2020. 8: p. 13937–13948.

13. Auer, P., N. Cesa-Bianchi, and P.J.M.L. Fischer, Finite-time Analysis of the Multiarmed Bandit Problem. 2004. 47: p. 235–256.

14. Li, C., et al., High dimensional Bayesian optimization using dropout, in Proceedings of the 26th International Joint Conference on Artificial Intelligence. 2017, AAAI Press: Melbourne, Australia. p. 2096–2102.

15. Adaptive platform trials: definition, design, conduct and reporting considerations. Nat Rev Drug Discov, 2019. 18(10): p. 797–807.

16. Thorlund, K., et al., Key design considerations for adaptive clinical trials: a primer for clinicians. 2018. 360: p. k698.

17. Allender, S., et al., Bayesian strategy selection identifies optimal solutions to complex problems using an example from GP prescribing. NPJ digital medicine, 2020. 3: p. 7–7.

18. Shiffman, S., A.A. Stone, and M.R. Hufford, Ecological momentary assessment. Annu Rev Clin Psychol, 2008. 4: p. 1–32.

19. Kessler, R. and D. Mroczek, An update of the development of mental health screening scales for the US National Health Interview Study. Ann Arbor: University of Michigan, Survey Research Center of the Institute for Social Research, 1992.

20. Andrews, G. and T. Slade, Interpreting scores on the Kessler Psychological Distress Scale (K10). Australian and New Zealand Journal of Public Health, 2001. 25(6): p. 494–497.

21. Australian Bureau of Statistics, Information Paper: Use of the Kessler Psychological Distress Scale in ABS Health Surveys, Australia, 2007-08. 2012, Commonwealth of Australia: Canberra, ACT, Australia.

22. Kessler, R.C., et al., Short screening scales to monitor population prevalences and trends in non-specific psychological distress. Psychol Med, 2002. 32(6): p. 959–76.

23. van Spijker, B.A.J., et al., The suicidal ideation attributes scale (SIDAS): Community-based validation study of a new scale for the measurement of suicidal ideation. Suicide and Life-Threatening Behavior, 2014. 44(4): p. 408–419.

24. Ford, K.L., et al., Youth Study Recruitment Using Paid Advertising on Instagram, Snapchat, and Facebook: Cross-Sectional Survey Study. JMIR Public Health Surveill, 2019. 5(4): p. e14080.

25. Guillory, J., et al., Recruiting Hard-to-Reach Populations for Survey Research: Using Facebook and Instagram Advertisements and In-Person Intercept in LGBT Bars and Nightclubs to Recruit LGBT Young Adults. J Med Internet Res, 2018. 20(6): p. e197.

26. Ellis, L.A., et al., Encouraging young men’s participation in mental health research and treatment: perspectives in our technological age. Clin Investig, 2014. 4(10): p. 881–888.

27. Sanchez, C., et al., Social media recruitment for mental health research: A systematic review. Compr Psychiatry, 2020. 103: p. 152197.

28. Kabat-Zinn, J., Mindfulness-based interventions in context: past, present, and future. Clinical psychology: Science and practice, 2003. 10(2): p. 144–156.

29. Cavanagh, K., et al., Can mindfulness and acceptance be learnt by self-help?: A systematic review and meta-analysis of mindfulness and acceptance-based self-help interventions. Clinical Psychology Review, 2014. 34(2): p. 118–129.

30. Sevinc, G., et al., Common and Dissociable Neural Activity After Mindfulness-Based Stress Reduction and Relaxation Response Programs. Psychosom Med, 2018. 80(5): p. 439–451.

31. Tang, Y.-Y., B.K. Hölzel, and M.I. Posner, The neuroscience of mindfulness meditation. Nature Reviews Neuroscience, 2015. 16(4): p. 213–225.

32. Klika, B. and C. Jordan, HIGH-INTENSITY CIRCUIT TRAINING USING BODY WEIGHT: Maximum Results With Minimal Investment. ACSM’s Health & Fitness Journal, 2013. 17(3).

33. American Academy of Sleep Medicine, International classification of sleep disorders. 2014.

34. Chung, K.F., et al., Sleep hygiene education as a treatment of insomnia: a systematic review and meta-analysis. Fam Pract, 2018. 35(4): p. 365–375.

35. Morin, C.M. and N.H. Azrin, Stimulus control and imagery training in treating sleep-maintenance insomnia. Journal of Consulting and Clinical Psychology, 1987. 55(2): p. 260–262.

36. Kang, Y.S., S.Y. Choi, and E. Ryu, The effectiveness of a stress coping program based on mindfulness meditation on the stress, anxiety, and depression experienced by nursing students in Korea. Nurse Educ Today, 2009. 29(5): p. 538–43.

37. Gee, B.L., et al., Suicidal thoughts, suicidal behaviours and self-harm in daily life: A systematic review of ecological momentary assessment studies. Digit Health, 2020. 6: p. 2055207620963958.

38. Broome, M.R., et al., Mood instability: significance, definition and measurement. Br J Psychiatry, 2015. 207(4): p. 283–5.

39. Beames, J.R., K. Kikas, and A. Werner-Seidler, Prevention and early intervention of depression in young people: an integrated narrative review of affective awareness and Ecological Momentary Assessment. BMC Psychology, 2021. 9(1): p. 113.

40. Dubad, M., et al., A systematic review of the psychometric properties, usability and clinical impacts of mobile mood-monitoring applications in young people. Psychol Med, 2018. 48(2): p. 208–228.

41. Shiffman, S., Commentary on McCarthy et al. (2015): Ecological momentary assessment--Reactivity? Intervention? Addiction, 2015. 110(10): p. 1561–2.

42. Antony, M.M., et al., Psychometric properties of the 42-item and 21-item versions of the Depression Anxiety Stress Scales in clinical groups and a community sample. Psychological assessment, 1998. 10(2): p. 176.

43. Shaw, T., et al., Properties of the DASS-21 in an Australian Community Adolescent Population. J Clin Psychol, 2017. 73(7): p. 879–892.

44. Greenwood, J.L., E.A. Joy, and J.B. Stanford, The Physical Activity Vital Sign: a primary care tool to guide counseling for obesity. Journal of Physical Activity and Health, 2010. 7(5): p. 571–576.

45. Golightly, Y.M., et al., Physical Activity as a Vital Sign: A Systematic Review. Prev Chronic Dis, 2017. 14: p. E123.

46. Buysse, D.J., et al., The Pittsburgh Sleep Quality Index: a new instrument for psychiatric practice and research. Psychiatry research, 1989. 28(2): p. 193–213.

47. Park, T., M. Reilly-Spong, and C.R. Gross, Mindfulness: a systematic review of instruments to measure an emergent patient-reported outcome (PRO). Quality of Life Research, 2013. 22(10): p. 2639–2659.

48. Baer, R., et al., Differential sensitivity of mindfulness questionnaires to change with treatment: A systematic review and meta-analysis. Psychological Assessment, 2019. 31(10): p. 1247–1263.

49. Baer, R., Assessment of mindfulness by self-report. Current Opinion in Psychology, 2019. 28: p. 42–48.

50. Thompson, E.R., Development and Validation of an Internationally Reliable Short-Form of the Positive and Negative Affect Schedule (PANAS). Journal of Cross-Cultural Psychology, 2007. 38(2): p. 227–242.

51. Tennant, R., et al., The Warwick-Edinburgh Mental Well-being Scale (WEMWBS): development and UK validation. Health and Quality of Life Outcomes, 2007. 5(1): p. 63.

52. Stewart-Brown, S., et al., Internal construct validity of the Warwick-Edinburgh mental well-being scale (WEMWBS): A rasch analysis using data from the Scottish health education population survey. Health Qual Life Outcomes, 2009. 7: p. 15.

53. Zimet, G.D., et al., The multidimensional scale of perceived social support. Journal of personality assessment, 1988. 52(1): p. 30–41.

54. Adler, N.E., et al., Relationship of subjective and objective social status with psychological and physiological functioning: preliminary data in healthy white women. Health Psychol, 2000. 19(6): p. 586–92.

55. National Institute on Drug Abuse, NIDA Drug Screening Tool, NIDA-Modified ASSIST (NM ASSIST). 2020, National Institutes of Health: Bethseda, MA, USA.

56. Smith, P.C., et al., A Single-Question Screening Test for Drug Use in Primary Care. Archives of Internal Medicine, 2010. 170(13): p. 1155–1160.

57. Devilly, G.J. and T.D. Borkovec, Psychometric properties of the credibility/expectancy questionnaire. J Behav Ther Exp Psychiatry, 2000. 31(2): p. 73–86.

58. Thompson-Hollands, J., et al., Credibility and outcome expectancy in the unified protocol: Relationship to outcomes. Journal of Experimental Psychopathology, 2014. 5(1): p. 72–82.

59. McConnaughy, E., J. Prochaska, and W. Velicer, Stages of change in psychotherapy: Measurement and sample profiles. Psychotherapy: Theory, Research & Practice, 1983. 20: p. 368–375.

60. National Institute for, H. and E. Care, NICE Process and Methods Guides, in Guide to the Methods of Technology Appraisal 2013. 2013, National Institute for Health and Care Excellence (NICE) Copyright © 2013 National Institute for Health and Clinical Excellence, unless otherwise stated. All rights reserved.: London.

61. Health, D.o., Procedure guidance for listing medicines on the Pharmacuetical Benefits Scheme (icluding consideration of vaccines for the National Immunisation Program) Version 1. 2016, Commonwealth of Australia: Canberra, ACT.

62. Committee, M.S.A., Technical guidelines for preparing assessment reports for the Medical Services Advisory Commitee-Medical Service Type: Therapeutic (Version 2.0), D.o. Health, Editor. 2016, Commonwealth of Australia.

63. EuroQol Research Foundation, EQ-5D-5L User Guide, Version 3.0. 2021.

64. Norman, R., P. Cronin, and R. Viney, A pilot discrete choice experiment to explore preferences for EQ-5D-5L health states. Appl Health Econ Health Policy, 2013. 11(3): p. 287–98.

65. Hunter, R.M., et al., An educational review of the statistical issues in analysing utility data for cost-utility analysis. Pharmacoeconomics, 2015. 33(4): p. 355–66.

66. Keetharuth, A.D., et al., Recovering Quality of Life (ReQoL): a new generic self-reported outcome measure for use with people experiencing mental health difficulties. The British journal of psychiatry : the journal of mental science, 2018. 212(1): p. 42–49.

67. Bouwmans, C., et al., The iMTA Productivity Cost Questionnaire: A Standardized Instrument for Measuring and Valuing Health-Related Productivity Losses. Value Health, 2015. 18(6): p. 753–8.

68. Brooke, J., SUS -- a quick and dirty usability scale. 1996. p. 189–194.

69. Zhou, L., et al., The mHealth App Usability Questionnaire (MAUQ): Development and Validation Study. JMIR Mhealth Uhealth, 2019. 7(4): p. e11500.

70. Lovibond, S.H. and P.F. Lovibond, Manual for the depression anxiety stress scales (2nd edition). 1995, Psychology Foundation: Sydney, NSW, Australia.

71. Carpenter and M.G. Kenward. Missing data in randomised controlled trials: a practical guide. 2007.

72. Satterthwaite, F.E., An Approximate Distribution of Estimates of Variance Components. Biometrics Bulletin, 1946. 2(6): p. 110–114.

73. Wilson, E.C., A practical guide to value of information analysis. Pharmacoeconomics, 2015. 33(2): p. 105–21.

74. Flight, L., et al., Expected Value of Sample Information to Guide the Design of Group Sequential Clinical Trials. Medical Decision Making, 2021. 42(4): p. 461–473.

75. Strong, M., et al., Estimating the Expected Value of Sample Information Using the Probabilistic Sensitivity Analysis Sample: A Fast, Nonparametric Regression-Based Method. Medical Decision Making, 2015. 35(5): p. 570–583.

76. Rothery, C., et al., Value of Information Analytical Methods: Report 2 of the ISPOR Value of Information Analysis Emerging Good Practices Task Force. Value in health : the journal of the International Society for Pharmacoeconomics and Outcomes Research, 2020. 23(3): p. 277–286.

77. Wang, S., D. Gum, and T. Merlin, Comparing the ICERs in Medicine Reimbursement Submissions to NICE and PBAC-Does the Presence of an Explicit Threshold Affect the ICER Proposed? Value Health, 2018. 21(8): p. 938–943.

78. Fenwick, E., et al., Value of Information Analysis for Research Decisions—An Introduction: Report 1 of the ISPOR Value of Information Analysis Emerging Good Practices Task Force. Value in Health, 2020. 23(2): p. 139–150.

79. National Health and Medical Research Council, A.R.C.a.U.A., Australian Code for the Responsible Conduct of Research. 2018, Commonweath of Australia: Canberra.

80. Australian Bureau of Statistics. Indigenous Status Standard. 2014; Available from: https://www.abs.gov.au/statistics/standards/indigenous-status-standard/latest-release#definition-of-variable.

81. Australian Bureau of Statistics. Ancestry Standard. 2014; Available from: https://www.abs.gov.au/statistics/standards/ancestry-standard/latest-release.

82. Brooke, J., SUS: a “quick and dirty” usability scale, in Usability evaluation in industry, P.W. Jordan, et al., Editors. 1996, Taylor & Francis Ltd.: London, UK. p. 189.

