## Supplementary information for "Protocol for a bandit-based response adaptive trial to evaluate the effectiveness of brief self-guided digital interventions for reducing psychological distress in university students: The Vibe Up Study"

#### Demographic details (DEM)

| | | | | | | |
|---|---|---|---|---|---|---|
| ☒ Screening | ☐ Baseline | ☐ EMA | ☐ Mid | ☐ Intervention | ☐ Post | ☐ Follow-up |
| | | | | | | Up to 10 questions |

*Source:* ABS Draft Standard for Sex, Gender, Variation of Sex Characteristics and Sexual Orientation Variables, 2020 (Unpublished), [80] and [81].

*Remarks:* DEM1b, DEM2b and DEM3b are included principally to afford (potentially marginalised) individuals the right to assert their identity.

| Id | Question | Presentation |
|---|---|---|
| DEM1a | How do you describe your gender?<br><br>This information will help our research understand if different groups of people find different treatments more useful. | 5-item closed choice: Woman or female (0), Man or male (1), Non-binary (2), I use a different term (3), Prefer not to say (4). User sees: words only. |

*If response to DEM1a is 3 (I use a different term):*

| | | |
|---|---|---|
| DEM1b | Please specify: | Free text, max 64 characters. |

*For all participants:*

| | | |
|---|---|---|
| DEM2a | What was your sex recorded at birth? | 3-item closed choice: Female (0), Male (1), Another term (2), Prefer not to say (3). User sees: words only. |

*If response to DEM2a is 2 (Another term):*

| | | |
|---|---|---|
| DEM2b | Please specify: | Free text, max 64 characters. |

*For all participants:*

| | | |
|---|---|---|
| DEM3a | How do you describe your sexual orientation?<br><br>This information will help our research understand if different groups of people find different treatments more useful. | 6-item closed choice: Straight (heterosexual) (0), Gay or lesbian (1), Bisexual (2), I use a different term (3), Don't know (4), Prefer not to say (5). User sees: words only. |

*If response to DEM3a is 3 (I use a different term):*

| | | |
|---|---|---|
| DEM3b | Please specify: | Free text, max 64 characters. |

*For all participants:*

| | | |
|---|---|---|
| DEM4 | Are you of Aboriginal or Torres Strait Islander origin? | 5-item multiple choice: No (0), Yes, Aboriginal (1), Yes, Torres |

| | | |
|---|---|---|
| | If you are of both Aboriginal and Torres Strait Islander origin, mark both 'Yes' boxes. | Strait Islander (2), Don't know (3), Prefer not to say (4). User sees: words only. Validation: If options 0, 3 or 4 are selected then no other option can be chosen. |

| | | |
|---|---|---|
| DEM5 | What is your ancestry? Pick up to two options. | 12-item multiple choice: Australian (0), Chinese or Cantonese (1), Dutch (2), English (3), German (4), Greek (5), Indian (6), Irish (7), Italian (8), Scottish (9), Vietnamese (10), Another ancestry (11), Prefer not to say (12). User sees: words only. Validation: Maximum two concurrent selections. If item 11 is chosen, no other items can be selected. |
| DEM6 | What is the language that you mostly speak at home? | 11-item closed choice: Arabic (0), Cantonese (1), English (2), Greek (3), Hindi (4), Italian (5), Mandarin (6), Punjabi (7), Spanish (8), Vietnamese (9), Another language (10), Prefer not to say (11). User sees: words only. |
| DEM7 | What is your postcode? | Numeric integer input, max 4 characters |

#### Study and employment (WRK)

| ☒ Screening | ☐ Baseline | ☐ EMA | ☐ Mid | ☐ Intervention | ☐ Post | ☐ Follow-up |
|---|---|---|---|---|---|---|
| | | | | | | Up to 3 questions |

*Source: Bespoke questions.*

| Id | Question | Presentation |
|---|---|---|
| WRK1 | Are you studying in Australia as an international student? | 2-item closed choice: No (0), Yes (1). User sees: words only. |
| WRK2a | What is your Weighted Average Mark (WAM) or Grade Point Average (GPA) for your **current** degree or qualification?<br><br>The WAM or GPA provides an indication of your overall academic performance. Please select the option that your institution uses. | 6-item closed choice with options: WAM (0), GPA – 4.0 scale (1), GPA – 7.0 scale (2), Don't know (3), Don't have one (4), Prefer not to say (5). User sees: numbers/words only. |

*If response to WRK2a is 0 (WAM):*

| Id | Question | Presentation |
|---|---|---|
| WRK2a | What is your Weighted Average Mark (WAM) for your **current** degree or qualification? | 11-item closed choice: Below 50 (0), 50-54 (1), 55-59 (2), 60-64 (3), 65-69 (4), 70-74 (5), 75-79 (6), 80-84 (7), 85-89 (8), 90-94 (9), 95 or higher (10), User sees: words only. |

*If response to WRK2a is 1 (GPA – 4.0 scale):*

| Id | Question | Presentation |
|---|---|---|
| WRK2a | What is your Grade Point Average (GPA – 4 point scale) for your **current** degree or qualification? | 8-item closed choice: Below 1.0 (0), 1.0-1.4 (1), 1.5-1.9 (2), 2.0-2.4 (3), 2.5-2.9 (4), 3.0-3.4 (5), 3.5-3.9 (5), 4.0 (7). User sees: numbers/words only. |

*If response to WRK2a is 2 (GPA – 7.0 scale):*

| Id | Question | Presentation |
|---|---|---|
| WRK2a | What is your Grade Point Average (GPA – 7 point scale) for your **current** degree or qualification? | 8-item closed choice: Below 4.0 (0), 4.0-4.4 (1), 4.5-4.9 (2) 5.0-5.4 (3), 5.5-5.9 (4), 6.0-6.4 (5), 6.5-6.9 (6), 7.0 (7). User sees: numbers/words only. |

#### Prior use of wellbeing strategies (PUWS)

| ☒ Screening | ☐ Baseline | ☐ EMA | ☐ Mid | ☐ Intervention | ☐ Post | ☐ Follow-up |
|---|---|---|---|---|---|---|
| | | | | | | 9 questions |

*Source: Bespoke questions.*

We would like to know more about your previous use of different wellbeing strategies.

| Id | Question | Presentation |
|---|---|---|
| PUWS1 | Have you ever used a smartphone app related to the following:<br><br>a. Mindfulness<br>b. Physical activity/exercise<br>c. Tips to improve your sleep | 2-item closed choice: No (0), Yes (1). User sees: words only. |
| PUWS2 | Have you ever taken part in a group program, online course or individual appointments related to the following:<br><br>a. Mindfulness<br>b. Physical activity/exercise<br>c. Tips to improve your sleep | 2-item closed choice: No (0), Yes (1). User sees: words only. |
| PUWS3 | How many days in the past week, have you:<br><br>a. Practised mindfulness<br>b. Engaged in physical activity/exercise<br>c. Used strategies to improve sleep | Numeric fill-in/pick 'X day(s)'. Validation: X is integral in the range $0 \leq X \leq 7$. |

### Daily log of engagement with intervention (LOG)

<table>
<tr><td>☐ Screening</td><td>☐ Baseline</td><td>☐ EMA</td><td>☐ Mid</td><td>☒ Intervention</td><td>☐ Post</td><td>☐ Follow-up</td></tr>
<tr><td colspan="7" align="right">1 question</td></tr>
</table>

*Source:* Bespoke questions.

*Remarks:* Participants will be encouraged to log their engagement with the intervention once per day during the intervention period. Participants will only see the option corresponding to the intervention they are allocated to (LOGa for Mindfulness, LOGb for Physical Activity, LOGc for Sleep Hygiene); the active control (EMA) condition will not see any question. Completion of the question is not compulsory.

<table>
<tr><th>Id</th><th>Question</th><th>Presentation</th></tr>
<tr><td>LOGa</td><td>How much time did you spend practicing mindfulness yesterday?</td><td rowspan="3">
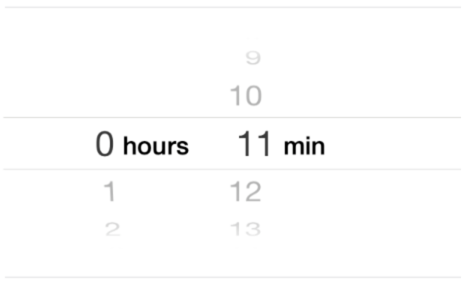

Timespan in hours and minutes.<br>Hours: 0 – 23 (coded 0 – 23).<br>Mins: 0 – 59 (coded 0 – 59)<br>Default: 0 hours and 0 minutes.</td></tr>
<tr><td>LOGb</td><td>How much time did you spend being physically active yesterday?<br><br>Please include any changes you made to increase your physical activity throughout the day (not just exercise workouts).</td></tr>
<tr><td>LOGc</td><td>How much sleep did you get last night?</td></tr>
</table>

*Scoring:* Report individual items only.

#### Within-study exposures questionnaire (EXP)

| ☐ Screening | ☐ Baseline | ☐ EMA | ☐ Mid | ☐ Intervention | ☒ Post | ☐ Follow-up |
|---|---|---|---|---|---|---|
| | | | | | | 4 questions |

*Source:* Bespoke questions.

*Remarks:* Rather than trying to enumerate exposures (which nevertheless may be evaluated differently by different people), the approach adopted here is to elicit *perception of perceived impact* on daily life, mental health or app use of any recent happening, regardless of cause.

| Id | Question | Measurement | Purpose |
|---|---|---|---|
| EXP1 | In the past **two weeks**, my life or routine was disrupted for some reason (ignoring anything to do with this app.) | 5-level Likert, Strongly disagree (1) – Disagree (2) – Undecided (3) – Agree (4) – Strongly agree (5), User sees: words only. | Life event confound |
| EXP2a | In the past **two weeks**, something **negatively** affected my mental health (ignoring anything to do with this app.) | | Mental health change confound |
| EXP2b | In the past **two weeks**, something **positively** affected my mental health (ignoring anything to do with this app.) | | Mental health change confound |
| EXP3 | In the past **two weeks**, something interfered with my ability to use this app. | | (Attitudinal) non-engagement confound |

*Scoring:* Report individual items only.

#### UX questionnaire (UX)

| ☐ Screening | ☐ Baseline | ☐ EMA | ☐ Mid | ☐ Intervention | ☒ Post | ☐ Follow-up |
|---|---|---|---|---|---|---|
| | | | | | | Up to 9 questions |

*Source:* Bespoke questions, based on the System Usability Scale[82] and mHealth App Usability Questionnaire[69].

| Id | Question | Measurement | Purpose |
|---|---|---|---|
| UX1 | I found the app easy to use. | 5-level Likert, Strongly disagree (1) – Disagree (2) – Undecided (3) – Agree (4) – Strongly agree (5), User sees: words only. | Usability: ease of use construct |
| UX2 | I found the app useful for my mental health. | | Usability: usefulness construct |
| UX3 | Overall, I am satisfied with the app. | | Usability: satisfaction construct |
| UX4a | I had no problems using the app. | | Technology barriers |

*If response to UX4A is 'Strongly Disagree' or 'Disagree'…*

| | | | |
|---|---|---|---|
| UX4b | Can you tell us what problem(s) you encountered?<br><br>Letting us know will help us improve the experience for others. Thank you! | Free text, Max 256 characters. Validation: non-null. | Technology barriers |

*For participants in an intervention arm (mindfulness, physical exercise, sleep hygiene) only…*

| | | | |
|---|---|---|---|
| UX5 | I trusted the information given by the app. | 5-level Likert, Strongly disagree (1) – Disagree (2) – Undecided (3) – Agree (4) – Strongly agree (5), User sees: words only. | Post-hoc credibility |
| UX6 | Activities or actions suggested by the app **were new** for me. | | Behaviour-change confound |
| UX7 | **In the past two weeks**, I have put into practice activities or actions suggested by the app. | | Subjective compliance |
| UX8 | I **intend in the future** to put into practice activities or actions suggested by the app. | | Post-hoc intention |

*Scoring:* Report individual items only.
